# Cardiovascular disease Multimorbidity is Associated with Decreased Health-Related Quality of Life in Haiti: Findings from a Population-based Cohort

**DOI:** 10.1101/2024.05.08.24307093

**Authors:** Shalom Sabwa, Vanessa Rouzier, Rodney Sufra, Reichling St. Sauveur, Nour Mourra, Rehana Rasul, Joseph Inddy, Lily D. Yan, Madeline Sterling, Laura Pinheiro, Marie Deschamps, Jean William Pape, Margaret L. McNairy

**Affiliations:** Department of Global Health and Population, Harvard T.H. Chan School of Public Health, Boston, Massachusetts, United States of America; Center for Global Health, Weill Cornell Medicine, 402 East 67th Street, New York, NY 10065, USA; Haitian Group for the Study of Kaposi’s Sarcoma and Opportunistic Infections (GHESKIO), 33 Boulevard Harry Truman, Port-au-Prince 6110, Haiti; Department of Epidemiology and Biostatistics, Graduate School of Public Health and Health Policy, City University of New York, New York, NY 10017, USA; Institute of Implementation Science in Population Health, City University of New York, New York, NY 10027, USA; Division of General Internal Medicine, Department of Medicine, Weill Cornell Medicine, 525 East 68th Street, Box 331, New York, NY, 10065, USA

**Author notes:** corresponding author: Shalom Sabwa, 665 Huntington Avenue, Room 1108, Boston, MA 02115.

## Abstract

**Background:** Multimorbidity is increasingly prevalent in lower-and middle-income countries (LMICs). Health-related quality of life (HRQOL) has been inversely associated with multimorbidity but is understudied in LMICs. We report cardiovascular disease (CVD) multimorbidity in Haiti and its association with HRQOL.

**Methods:** We used data from the Haiti CVD Cohort, a population-based longitudinal cohort of adults. CVD multimorbidity was 2+ CVD risk factors/diseases at enrollment. HRQOL was measured using the Short Form-12, yielding physical (PCS)/mental (MCS) component summary scores between 0-100, with higher scores indicating better HRQOL. We used linear regressions to assess the association between CVD multimorbidity and HRQOL and between individual CVD comorbidities and HRQOL. Additionally, we examined how gender and education modified the main effect.

**Results:** Among 2,996 participants, the median age was 40 (IQR: 27-55), 58.0% were female, and 70.3% earned <1 USD/day. CVD multimorbidity prevalence was 24.1%; compared to those without CVD multimorbidity, those with CVD multimorbidity were older (median 56.0 [IQR: 47.0, 53.0]) and female (70.5%). Adjusted models revealed CVD multimorbidity was inversely related to PCS (−2.7 [95% CI: −3.8, −1.6]) and MCS (−1.0 [95% CI: −1.8, −0.2]). Heart failure and hypertension showed the strongest CVD morbidities associated with poor HRQOL. In the interaction analysis, among men, CVD multimorbidity was associated with a 4.3-point lower PCS score, and among those with less education, CVD multimorbidity was associated with a 4.6-point lower PCS score than no CVD multimorbidity.

**Conclusions:** Our data are among the first to describe HRQOL data with high CVD multimorbidity in a young population in urban Haiti, and CVD multimorbidity was associated with decreased HRQOL.

(https://www.clinicaltrials.gov/ct2/show/NCT03892265, #NCT03892265)

**Research Perspective:**

– This research raises the question of whether people with CVD multimorbidity in LMICs need specific interventions that are tailored to their high levels of comorbidity and if women and those with low socioeconomic status are at extra risk for experiencing the adverse effects of multimorbidity.
– Future research should investigate cut-off points for describing a population average relevant to an LMIC.

## Introduction

Cardiovascular disease (CVD) is the leading cause of death globally, with 80% of the burden in lower- and middle-income countries (LMICs).^1,2^ Haiti is the first black republic in the world and a nation of remarkable resilience, given decades of natural disasters, political turmoil, and economic instability.^3^ Over the last two decades, CVD is estimated to have surpassed HIV in Haiti as the leading cause of mortality, and modeling data suggest CVD accounted for 36% of adult deaths in 2019.^4,5^ Prior research in Haiti has also found high rates of early-onset CVD risk factors, including hypertension, obesity, and renal disease.^6–8^

Multimorbidity is the presence of two or more chronic diseases and is often the result of non-communicable diseases (NCDs) sharing similar biological risk factors and social determinants of health.^9^ Individuals with multimorbidity are at a greater risk for disability and premature mortality.^10–12^ Persons with multimorbidity are also associated with greater use of healthcare service resources and unplanned hospital admissions.^13^ Reliable population-based estimates of CVD multimorbidity in LMICs are limited, given the lack of local population-based epidemiologic data. Assessing the prevalence and impact of CVD multimorbidity is essential to understanding the burden of CVD in LMICs and to guide future screening and treatment.

In addition to its associated morbidity and mortality, multimorbidity has been inversely associated with adverse health-related quality of life (HRQOL) in high-income settings.^12,14–16^ HRQOL instruments assess how well an individual believes they can function daily and their perception of their physical, mental, and social well-being.^17^ HRQOL has become an increasingly crucial patient-centered factor due to its ability to predict adherence to a medical regimen, healthcare utilization, CVD events, and overall mortality.^18,19^ HRQOL metrics may help identify individuals who can benefit the most from increased screenings and interventions.^14,18^

Prior research has already demonstrated that patients with CVD experience a lower quality of life than those without CVD in developed countries.^17^ However, there are knowledge gaps on the impact of multiple CVD risk factors and cardiovascular diseases on HRQOL in adults in non-European or Western cohorts and severely impoverished settings.^18–20^ This research would help contextualize how experiencing numerous chronic diseases truly impacts the quality of life in LMICs and can inform future interventions to address HRQOL among those with CVD.

To address this gap, the primary goal of this study was to assess the prevalence of CVD multimorbidity and its association with HRQOL in a contemporary Haitian Cohort. In addition, we explored the impact of individual CVD diseases and risk factors on HRQOL and the interaction of gender and education on the relationship between CVD multimorbidity and HRQOL.

This study reports cross-sectional data from the Haiti CVD Disease Cohort (https://www.clinicaltrials.gov/ct2/show/NCT03892265, #NCT03892265), which is a population-based CVD cohort of Haitian adults living in urban Port-au-Prince, Haiti.

## Methods

### Study setting and design

Haiti is located on the western third of the island of Hispaniola in the Caribbean Sea. It has a population of 11 million, with 59% living in urban areas, including Port-au-Prince, Haiti’s capital city.^21^ This is a cross-sectional sub-analysis within the Haiti CVD Cohort Study, a longitudinal cohort study of adults in Port-au-Prince. Details of study procedures have been previously described.^22^

A total of 3,005 participants were enrolled in the parent cohort using multi-stage sampling between March 19, 2019, and August 23, 2021, at Groupe Haitien d’Etude de Sarcome de Kaposi et de Infections Opportunistes (GHESKIO), a clinic providing care, training, and conducting research in downtown Port-au-Prince. Participants were eligible for the study if they were ≥18 years old, had a primary residence in Port-au-Prince, and had no serious medical condition or cognitive impairment preventing participation at baseline.

This analysis evaluates cross-sectional data collected from participants in the Haiti CVD Cohort at study enrollment.

### Primary Outcome

The primary outcome of this analysis was HRQOL as measured by the Short-Form-12 (SF-12). The SF-12, a 12-item questionnaire developed by the Medical Outcomes Study, is a generic, psychometrically validated HRQOL instrument.^23^ It has been used extensively to capture HRQOL for individuals with chronic conditions. The SF-12 includes the Physical Component Summary (PCS) and Mental Health Component Summary (MCS) scores. The PCS and MCS each range from 0 to 100, with higher scores indicating better HRQOL. The minimum clinically important difference (MCID) is a change of five units for the PCS and the MCS. For this study, participants were included in this analysis if they had complete data on the SF-12. We operationalized PCS and MCS scores as continuous outcomes.

### Key Exposure CVD Multimorbidity

CVD multimorbidity was ascertained as 10 CVD-related risk factors and diseases diagnosed by physicians and reported in the Haiti CVD Cohort Study (Supplemental Table 1). CVD risk factors include diabetes, hyperlipidemia, obesity, kidney disease, and hypertension. The cardiovascular diseases included angina/myocardial infarction, heart failure, and stroke/transient ischemic attack (TIA). CVD diseases were adjudicated by a physician committee using the American Heart Association^24^, European Society of Cardiology^25^, and World Health Organization^26^ guidelines. This analysis defined CVD multimorbidity as two or more CVD risk factors or diseases assessed at enrollment. CVD multimorbidity was operationalized as a categorical variable with the following levels: no CVD comorbidities (zero CVD comorbidities), one CVD comorbidity, and two or more CVD comorbidities (also referred to as CVD Multimorbidity). If a participant had any missing data to diagnose a risk factor, the individual was considered not to have that risk factor or disease.

### Measurements

Data collected at study enrollment included socio-demographic information, health behaviors (i.e., physical activity, smoking, alcohol use, insert others), HRQOL using SF-12 survey, physiologic measures including blood pressure, physical exams, assessment for CVD symptoms and events, laboratory measures, electrocardiogram (ECG), and echocardiogram (ECHO).

### Covariates

Covariates were chosen based on similar studies on multimorbidity and HRQOL.^12,14^ Age was categorized into 10-year intervals from 18-59 and a final category of 60+. Gender was reported as male and female. Education level was categorized as completing primary or less, secondary education, and higher than secondary education. Household income was asked based on Haitian currency, then converted to U.S. dollars (USD) for comparability (one USD = roughly 90 Haitian gourdes during the study period). We further categorized income into three categories: equivalent to less than one USD/day (including those reporting no income), one to ten USD, and greater than 10 USD/day. Smoking status was categorized as lifetime use (yes/no) and current use (yes/no). Physical activity was categorized into less than or equal to 150 minutes per week and greater than 150 minutes weekly. Alcohol was dichotomized into low (less than or equal to one drink a day) and moderate-high use (greater than one drink daily). We examined cohort characteristics overall and by CVD morbidity burden category.

### Statistical Analysis

Descriptive statistics were used to summarize the cohort’s characteristics, including CVD multimorbidity. We assessed unadjusted differences in cohort characteristics across CVD multimorbidity groups using ANOVA or Chi-square tests. The ordinary least squares regression for multimorbidity was estimated for PCS and MCS subscales. Next, we used linear regression models to evaluate the association between CVD multimorbidity and SF-12 component summary scores (PCS and MCS), adjusted models for age, sex, education level, employment, and household income. Linear regression models were also used to assess the relative impact of each disease or risk factor on the PCS and MCS, adjusted for age, sex, education, income, smoking status, alcohol intake, and physical activity. Linear regression was also used to examine the potential for effect modification of the relationship between CVD and HRQOL due to gender and education. These covariates were chosen as potential effect modifiers based on health disparities for CVD risk for women, compared to men, and for those with low socioeconomic status. Missing data on covariates was less than one percent, and all linear regression models used a complete case analysis. Statistical significance was determined based on a *P* value < 0.05, and 95% confidence intervals were provided. All data analyses were performed in R.

### Ethics

The Weill Cornell Medicine and GHESKIO institutional review boards approved the study protocol and ethical consent forms. Before enrollment, individuals selected for the study provided written informed consent.

## Results

### Baseline characteristics of study participants

Of the 3,005 Haiti CVD Cohort participants, nine individuals were excluded based on missing values on the SF-12. Our final sample included 2,996 participants (99.7% of the total sample) with complete data on the SF-12. The baseline characteristics of the study participants are outlined in Table 1. Participants had a median age of 40 (interquartile range (IQR): 27, 55), with 48.4% under 40 years. Most of the participants in the cohort were female (58.0%). 35.8% of participants had no or only primary-level schooling, and 70.3% reported not working or making less than one USD/day. There was a low prevalence of tobacco and alcohol use, with 3.6% reporting current tobacco use, 7.4% reporting lifetime tobacco use, and 3.7% moderate-high alcohol use. Over half of the participants reported physical activity of less than 150 minutes weekly (53.3%).

**Table 1.** Socio-demographics and health behaviors for adults in the Haiti CVD Cohort stratified by the CVD Multimorbidity group.

|  | <u>Total</u><br><u>N (%)</u><br><u>N=2,996</u> | <u>No CVD</u><br><u>morbidity N (%)</u><br><u>N=1,505 (50.2)</u> | <u>1 CVD</u><br><u>morbidity</u><br><u>N (%)</u><br><u>N=769 (25.7)</u> | <u>2 or more CVD</u><br><u>morbidity</u><br><u>N (%)</u><br><u>N=722 (24.1)</u> |
| --- | --- | --- | --- | --- |
| **Age, years, Median [IQR]** | 40.0 [27.0, 55.0] | 30.0 [24.0, 42.0] | 45.0 [35.0, 57.0] | 56.0 [47.0, 53.0] |
| 18-29 | 885 (29.5) | 728 (48.4) | 126 (16.4) | 31 (4.3) |
| 30-39 | 567 (18.9) | 352 (23.4) | 156 (20.3) | 59 (8.2) |
| 40-49 | 532 (17.8) | 214 (14.2) | 178 (23.1) | 140 (19.4) |
| 50-59 | 498 (16.6) | 134 (8.9) | 154 (20.0) | 210 (29.1) |
| 60+ | 514 (17.2) | 55 (5.1) | 155 (20.2) | 282 (39.1) |
| **Sex** |  |  |  |  |
| Male | 1,258 (42.0) | 761 (51.1) | 276 (35.9) | 213 (29.5) |
| Female | 1,738 (58.0) | 736 (48.9) | 493 (64.1) | 509 (70.5) |
| **Education** |  |  |  |  |
| Primary or less | 1,073 (35.8) | 277 (18.4) | 342 (44.5) | 454 (62.9) |
| Secondary | 1,478 (49.3) | 898 (59.7) | 343 (44.6) | 237 (32.8) |
| Higher than Secondary | 445 (14.9) | 330 (21.9) | 84 (10.9) | 31 (4.3) |
| **Household Income (daily)** |  |  |  |  |
| No report of paid work/ <1 USD | 2,105 (70.3) | 1,601 (70.5) | 528 (68.7) | 516 (71.5) |
| 1-10 USD | 360 (12.0) | 172 (11.4) | 111 (14.4) | 77 (10.7) |
| >10 USD | 531 (17.7) | 272 (18.1) | 130 (16.9) | 129 (17.9) |
| **Smoking (current use)** |  |  |  |  |
| Current use of tobacco | 107 (3.6) | 54 (3.6) | 23 (3.0) | 30 (4.2) |
| No current use of tobacco | 2,886 (96.3) | 1,449 (96.3) | 745 (96.9) | 692 (95.8) |
| Missing | 3 (0.1) | 2 (0.1) | 1 (0.1) | 0 (0.0) |

|  | <u>Total</u><br><u>N (%)</u><br><u>N=2,996</u> | <u>No CVD</u><br><u>morbidity N (%)</u><br><u>N=1,505 (50.2)</u> | <u>1 CVD</u><br><u>morbidity</u><br><u>N (%)</u><br><u>N=769 (25.7)</u> | <u>2 or more CVD</u><br><u>multimorbidity</u><br><u>N (%)</u><br><u>N=722 (24.1)</u> |
| --- | --- | --- | --- | --- |
| **Smoking (lifetime use)** |  |  |  |  |
| Lifetime use of tobacco | 221 (7.4) | 135 (9.0) | 45 (5.9) | 41 (5.7) |
| No lifetime use | 2,763 (92.2) | 1,364 (90.6) | 721 (93.8) | 678 (93.9) |
| Missing | 12 (0.4) | 6 (0.4) | 3 (0.4) | 3 (0.4) |
| **Physical Activity** |  |  |  |  |
| ≤150 min/week | 1,594 (53.3) | 774 (51.5) | 417 (54.4) | 403 (56.0) |
| >150 min/week | 1,397 (46.7) | 730 (48.5) | 350 (45.6) | 317 (44.0) |
| Missing | 5 (0.2) | 0 (0.0) | 2 (0.3) | 2 (0.3) |
| **Alcohol** |  |  |  |  |
| Low (≤1 drink a day) | 2,876 (96.3) | 1,425 (94.9) | 746 (97.1) | 705 (98.2) |
| Moderate-high (> 1 drink a day) | 112 (3.7) | 77 (5.1) | 22 (2.9) | 13 (1.8) |
| Missing | 8 (0.3) | 3 (0.2) | 1 (0.1) | 4 (0.6) |

\* CVD=cardiovascular disease; IQR=interquartile range.
† CVD morbidity is based on 10 CVD risk factors and diseases.

### CVD Multimorbidity

Among 2,996 participants, 30.5% had hypertension, 17.1% had obesity, 11.7% had heart failure, 8.8% had kidney disease, 12.4% had hyperlipidemia, 5.4% had diabetes, 2.6% had stroke or TIA, and 2.4% had angina or MI.

Overall, 50.2% of participants had no CVD comorbidities, 25.7% had one CVD comorbidity, and 24.1% had CVD multimorbidity. Participants with CVD multimorbidity were older than those with no CVD comorbidities (median age 56.0 vs. 30.0 years). Compared to no CVD comorbidities, participants with CVD multimorbidity were also more likely to be women (70.5% vs. 48.9%) and have primary education or less (62.9% vs. 18.4%).

Among those with CVD multimorbidity, the most prevalent CVD-related chronic conditions were hypertension (82.8%), then obesity (42.1%), followed by heart failure (41.6%), kidney disease (28.6%), hyperlipidemia (37.8%), diabetes (18.7%), stroke (10.1%), and angina/MI (7.6%) (Figure 1).

**Figure 1:**
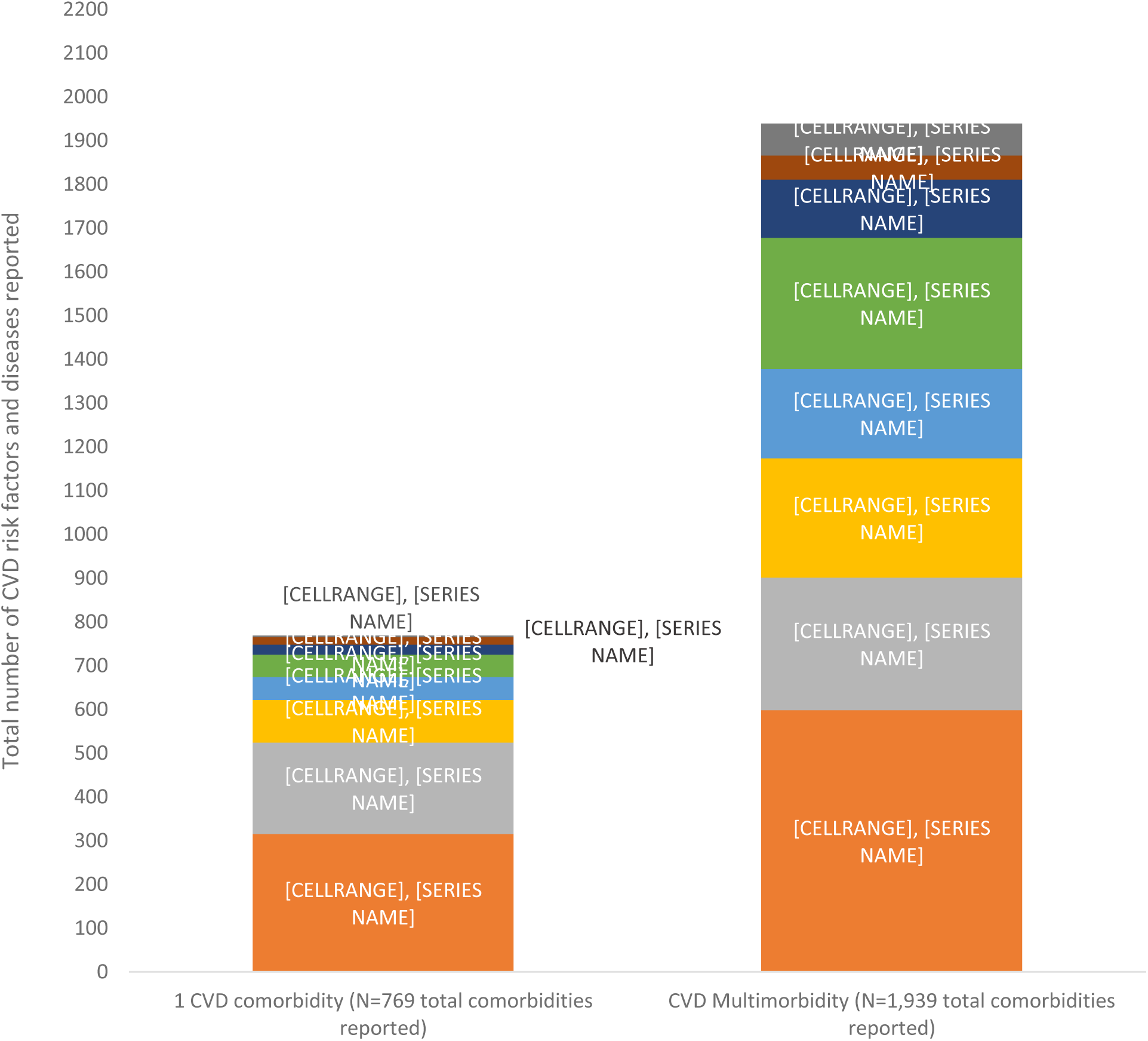
CVD risk factors and diseases within the CVD multimorbidity groups. * Participants with CVD multimorbidity had a combination of one or more of these conditions.

### HRQOL Among Those with CVD Multimorbidity

Overall, the mean PCS score was 37.0 (Standard deviation (SD): 11.4), and the mean MCS score was 46.8 (SD: 7.9). As the number of CVD comorbidities increased, the average PCS score decreased, and the average MCS score for the no comorbidities and 1 comorbidities group were similar, but those with CVD multimorbidity had the lowest MCS score (Figure 2). The mean PCS score for the three CVD morbidity groups was 39.6 for those without CVD comorbidities, 36.0 for those with one CVD comorbidity, and 32.5 for those with CVD multimorbidity (p-value <0.001) (Supplemental Table S3). The mean MCS score for the three CVD comorbidity groups was 46.9 for those without CVD morbidities, 47.2 for those with one CVD morbidity, and 46.0 for those with CVD multimorbidity (p value=0.00883). More significant differences between the CVD comorbidity groups were seen for the PCS scores compared to the MCS scores. The differences between the CVD comorbidity group scores for both models were statistically different, but only the PCS model had values more than 5 points different (the MCID for this study).

**Figure 2:**
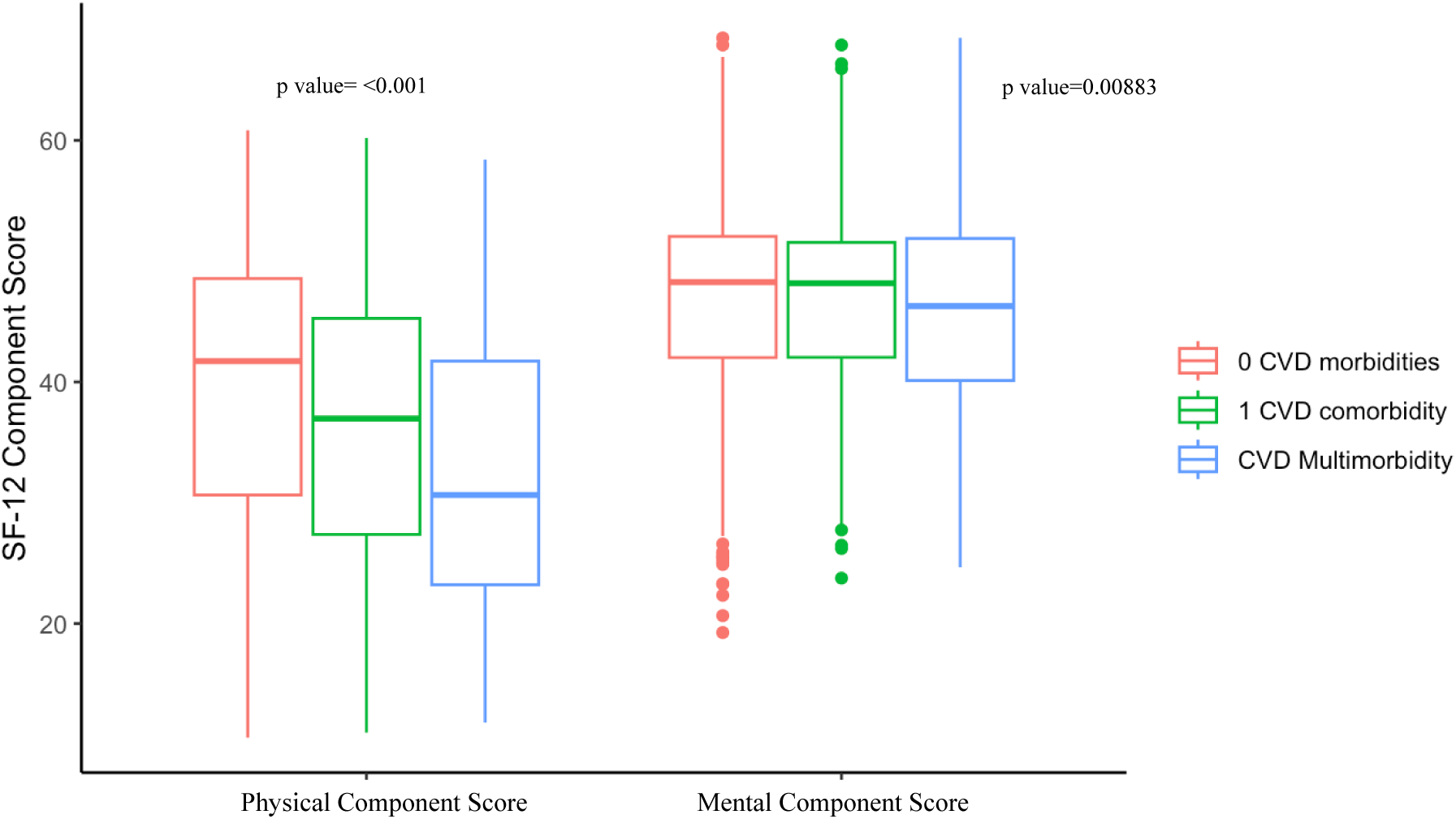
HRQOL measured by SF-12 Component Scores by CVD Multimorbidity Groups. * PCS and MCS scores based on Short-Form 12 score for each CVD multimorbidity group † Multimorbidity based on 10 CVD risk factors and diseases ‡ *p* values correspond with ANOVA for continuous variables that are normally distributed

### Association between CVD multimorbidity and MCS and PCS

We observed an inverse relationship between CVD multimorbidity and physical HRQOL (PCS) after adjusting for age, sex, education, income, smoking status, alcohol intake, and physical activity. Table 2 presents the results from the adjusted linear regression. On average, participants with CVD multimorbidity had PCS scores that were 2.7 points lower (95% CI: [-3.8,

**Table 2:** Linear regression models for CVD morbidity associated with HRQOL scores.

|  | PCS coefficient | MCS coefficient |
| --- | --- | --- |
| **Unadjusted model ($\beta$) [95% CI]** |  |  |
| No CVD comorbidity | Ref group | Ref group |
| 1 CVD comorbidity | -3.6 [-4.6, -2.7] | 0.2 [-0.5, 0.9] |
| CVD multimorbidity | -7.1 [-8.1, -6.1] | -1.0 [-1.7, -0.3] |
| **Adjusted model ($\beta$) [95% CI]** |  |  |
| No CVD comorbidity | Ref group | Ref group |
| 1 CVD comorbidity | -0.9 [-1.9, 0.0] | 0.1 [-0.6, 0.8] |
| CVD multimorbidity | -2.7 [-3.8, -1.6] | -1.0 [-1.8, -0.2] |

*\*Linear regression after adjusting for age, sex, education, income, smoking status, alcohol intake, and physical activity*

-1.6]) than those without CVD comorbidities. On average, participants with CVD multimorbidity had MCS scores that were 1.0 points lower [95% CI: −1.8, −0.2] than those without CVD comorbidity.

### Effect Modification by Gender and Education

Table 3 shows the results of the main effect with an interaction term between CVD multimorbidity and sex. Among males, CVD multimorbidity was associated with a PCS score that was 4.3 points lower (95% CI: [-7.2, −1.5]) compared to those with no CVD comorbidities. This association was not as strong among females (−2.2 points [95% CI: −4.7, 0.3]). Lastly, among participants with primary or less education, those with CVD multimorbidity had, on average, a PCS score 4.6 points lower (95% CI: [-7.2, −2.0]) than those with no CVD comorbidities.

**Table 3.** Linear regression models for the associations between CVD multimorbidity and HRQOL scores modified by sex and education level.

|  | PCS coefficient<br>[95% CI] | MCS coefficient<br>[95% CI] |
| --- | --- | --- |
| Among Females |  |  |
| 1 CVD comorbidity vs No CVD comorbidity | -0.3 (-2.3, 1.6) | 0.2 (-1.2, 1.7) |
| CVD multimorbidity vs No CVD comorbidity | -2.2 (-4.7, 0.3) | -1.7 (-3.5, 0.1) |
| Among Males |  |  |
| 1 CVD comorbidity vs No CVD comorbidity | -2.4 (-4.7, -0.1) | 0.1 (-1.6, 1.8) |
| CVD multimorbidity vs No CVD comorbidity | -4.3 (-7.2, -1.5) | 1.0 (-3.1, 1.1) |
| Among men and women with primary or less education |  |  |
| 1 CVD comorbidity vs No CVD comorbidity | -3.4 (-6.1, -0.7) | 0.6 (-1.4, 2.6) |
| CVD multimorbidity vs No CVD comorbidity | -4.6 (-7.2, -2.0) | -0.2 (-2.1, 1.7) |
| Among men and women with secondary education |  |  |
| 1 CVD comorbidity vs No CVD comorbidity | 0.2 (-1.9, 2.3) | -0.1 (-1.7, 1.5) |
| CVD multimorbidity vs No CVD comorbidity | -2.0 (-4.5, 0.6) | -1.3 (-3.2, 0.6) |
| Among men and women with higher than secondary education |  |  |
| 1 CVD comorbidity vs No CVD comorbidity | -0.9 (-4.9, 3.0) | -0.0 (-2.9, 2.9) |
| CVD multimorbidity vs No CVD comorbidity | -3.2 (-9.3, 2.9) | -2.5 (-7.0, 2.0) |

\* Interaction analyses of sex and education level after adjusting for age, income, smoking, alcohol, and physical activity.

Figure 3 shows the association between individual and combination diseases and the PCS and MCS. After adjusting for age, sex, education, income, smoking status, alcohol intake, and physical activity, hypertension was associated with a PCS score 1.3 points lower [95%CI: −2.2, −0.3], heart failure was associated with a PCS score 4.6 points lower [95%CI: −5.8, −3.4], stroke was associated with a PCS score 2.6 points lower [95% CI: −5.0, −0.2], kidney disease was associated with a PCS score 2.2 points lower, diabetes was associated with a PCS score 2.1 points lower [95%CI: −3.8, −0.4], the combination of hypertension and heart failure was associated with a PCS score 4.0 points lower [95%CI: −5.4, −2.6], and the combination of obesity and hypertension was associated with a PCS score 1.6 points lower [95%CI: −3.1, −0.2]. After adjusting for age, sex, education, income, smoking status, alcohol intake, and physical activity, angina was associated with an MCS score 3.5 points lower [95% CI: −5.3, −1.7], heart failure was associated with an MCS score 1.8 points lower [95% CI: −2.6, −0.9], stroke was associated with an MCS score 2.0 points lower [95%CI: −3.8, −0.3], the combination of hypertension and heart failure was associated with an MCS score 2.0 points lower [95%CI: −3.0, −0.9], and the combination of stroke and hypertension was associated with an MCS score 1.9 points lower [95%CI: −3.8, −0.1].

**Figure 3.**
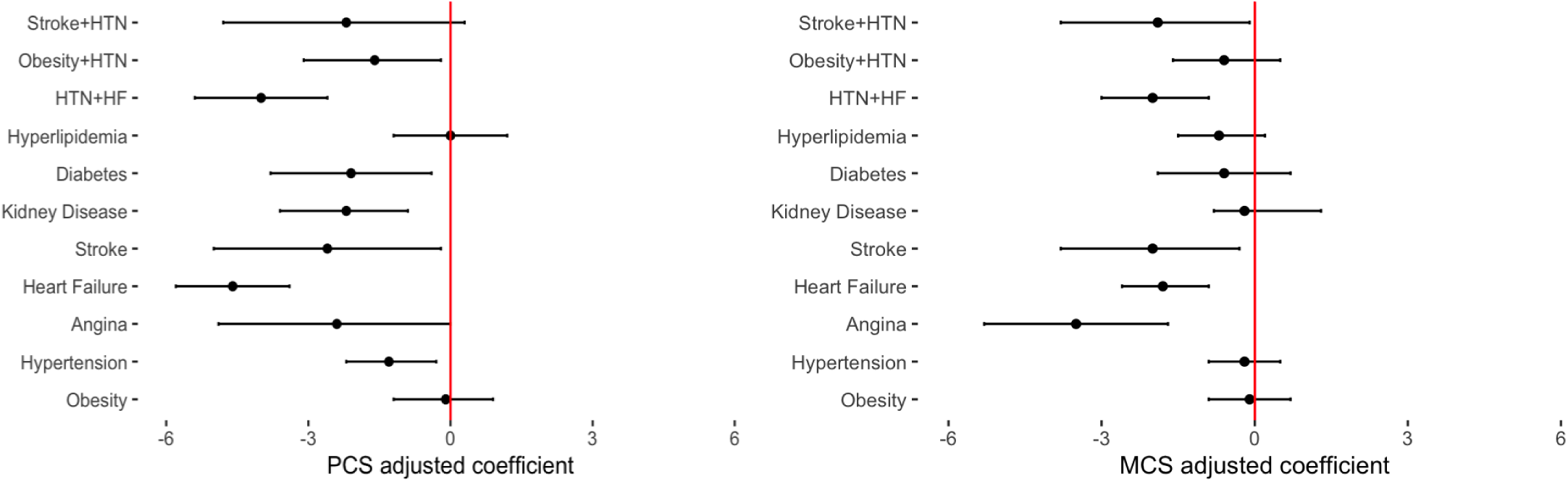
Adjusted estimates of changes to PCS and MCS scores (with 95%CI) based on regression models of individual CVD risk factors and disease categories. * Bars denote 95%CI, and dots represent the estimate. † Lines to the left of 0 indicate a decrease in HRQOL, and lines to the right indicate an increase.

## Discussion

This analysis quantifies the relationship between CVD multimorbidity and HRQOL among a cohort of adults living in an extremely low-income setting faced with an escalating CVD epidemic. A considerable proportion of adults, 24%, have CVD multimorbidity, with higher rates in older adults and females. High rates of hypertension and early-onset heart failure primarily drive CVD multimorbidity. Haitian self-reported health-related quality of life (HRQOL) is much lower than that of the US general population mean of 50, with an average PCS score of 37.0 and an average MCS score of 46.8. CVD multimorbidity in Haiti is associated with poorer HRQOL, particularly for physical health, compared to mental health, signifying coping mechanisms and psychological factors that highlight the population’s resilience despite challenging health conditions. On average, the study population had an MCS score 10 points higher than their PCS score. This difference is consistent with the literature on the impact of chronic diseases on health and highlights the dual effect of CVD multimorbidity.^16,27^

We found that individuals with hypertension and heart failure had considerably lower scores on the PCS and MCS than any other individual CVD risk factor or disease. Hypertension’s association with lower HRQOL is well-documented, and this study is consistent with the literature.^28,29^ In our study, individuals with hypertension had a 1.3-point lower score on the PCS. However, heart failure had the most significant impact on both HRQOL dimensions in our study. People with heart failure had PCS scores 4.6 points lower and MCS scores 1.8 points lower. Other studies have also documented that participants with heart failure generally have a substantial disease burden, and those who have additional comorbidities suffer from the worst prognosis and overall increased mortality.^14,30^ The impact of hypertension and heart failure on reducing quality of life is consistent with similar studies from different settings around the world.^29,31^ However, the magnitude of the association in this study is higher than most of these studies, as shown in a systematic review of hypertension and HRQOL by Trevisol et al.^29^ This implies that individuals experiencing the same clinical symptoms may feel significantly worse. This could be due to poverty, psychosocial stress, and political unrest in Port-au-Prince. Hypertension and heart failure have also previously been individually associated with depression.^32–34^ People with hypertension or heart failure likely suffer from both physical and mental health impairments, likely due to the stress of managing chronic conditions, suggesting that these patients require extra care and monitoring to alleviate their comorbidities. These patients may benefit from additional screening for mental health concerns, including depression, social, and family support. These patients may also be amenable to future interventions that target their complex disease treatment and management to improve their quality of life.

We found women had higher rates of CVD multimorbidity, which has been found in other studies.^1,35^ This may be due to social factors that expose women to risk factors different from those of men.^36^ For example, low economic and educational attainment status can influence a woman’s vulnerability to chronic diseases.^36,37^ In Haiti, women are generally less likely to have formal work arrangements and tend primarily to the home.^38^ This may increase their vulnerability to risk factors like physical inactivity and lack of income.^38,39^ To illustrate this point, Dade et al. reported a high rate of obesity in women in Haiti and suggested that social norms could substantially affect women’s health.^37^

This study contributes to the literature by reporting that gender is statistically significant for this cohort in affecting how one reports quality of life.^40^ While women were more likely to have CVD multimorbidity, the magnitude of the association between CVD multimorbidity and HRQOL was the greatest in men. For men with CVD multimorbidity, we found that their HRQOL was significantly decreased than those with no CVD comorbidities. Societal expectations and gender roles can influence how men and women perceive and express their health concerns. Men also tend to have higher rates of “acquired” risks such as drinking, smoking, and physical inactivity or risks in job hazards or recreational activities (such as strenuous leisure activities, i.e., exercising with heavy equipment) that affect their quality of life more severely than women.^39^ In addition, when men experience CVD multimorbidity, it may be more advanced or significantly impact their health due to delayed detection and intervention. Conversely, women may have more social resilience from social networks and support systems to mitigate the impact of CVD multimorbidity on self-reported HRQOL.^41–43^

Our study also found individuals with no education and multiple CVD comorbidities exhibited notably lower scores on the PCS than those without concurrent CVD comorbidities. We used low education as a proxy for low socioeconomic status, aligning with our prior work and other analyses.^44^ Prior investigations have consistently underscored the inverse relationship between lower education attainment and diminished HRQOL.^27,45,46^ This trend in our analysis persists even as one’s educational level rises. However, among individuals who completed at least secondary education or higher, the extent of the association between CVD multimorbidity and PCS diminishes. This implies that higher education might play a mitigating role in alleviating the adverse HRQOL effects associated with CVD multimorbidity, and longitudinal data is needed to establish causal inference.

A strength of this study is that the data comes from a population-based cohort, with adjudicated CVD events and definitions of risk factors comparable to US and international cohorts and SF-12 data. The study uniquely includes robust behavioral health questionaries and clinical data, including cardiac laboratory and imaging data. In addition, the study used a minimum clinically important difference to determine relevant changes. This study also contributes data that can be referenced as future research studies select cut-off points for describing a population average relevant to an LMIC. Subsequent research should incorporate qualitative analysis to better understand these patients’ perceptions regarding their compromised physical health. Limitations include the cross-sectional data analysis, which cannot establish a causal relationship between CVD multimorbidity and HRQOL. Data may not be generalizable to non-urban LMICs and do not include pregnancy.

Our findings underscore a high prevalence of CVD multimorbidity in Haiti, with the burden greatest in women compared to men. CVD multimorbidity is associated with decreased HRQOL, which has a larger effect on physical health-related quality of life. These findings support the importance of enhancing CVD prevention, especially among women, and screening for the impact of heart disease on patient’s mental and physical quality of life.

## Data Availability

The datasets used and/or analyzed during the current study are available from the corresponding author on reasonable request. Data request should be submitted to the study PI, Dr. Margaret McNairy who will review the data request with Haiti GHESKIO Site PI, Dr. Jean Pape and the study?s Observational Monitoring Board for approval.

## Acknowledgments

None

## Non-standard Abbreviations and Acronyms

SF-12: Short form 12
PCS: Physical component summary
MCS: Mental component summary

